# Characterizing patient details-related challenges from health information technology-related incident reports from Swedish healthcare

**DOI:** 10.1101/2023.05.30.23290728

**Authors:** Md Shafiqur Rahman Jabin, Ding Pan, Evalill Nilsson

## Abstract

This study examined health information technology-related (HIT) incidents to identify patient details-related issues, their association with contributing factors, and outcomes. Sources of information comprised retrospectively collected incident reports (n=95) using two sampling methods, i.e., purposive and snowball sampling. The reports were collected in two formats – interviews (written and telephone) and/or a set of already existing reports from the local database. The incident reports were analyzed using both the inductive method (thematic analysis) and the deductive approach using an existing framework, i.e., the International Classification for Patient Safety. The studies identified 90 incidents with 120 patient details-related issues—categorized as either information-related (48%) or documentation-related (52%) problems; around two-thirds of the 120 issues were characterized by human factors. Of the total sample, 87 contributing factors were identified, of which “medical device/system” (45%) and “documentation” (20%) were the most common contributing factors. Of 90 incidents, more than half (59%) comprised patient-related outcomes—patient inconvenience (47%) and patient harm (12%) and the remaining 41% (n=37) included staff or organization-related outcomes. The study confirms that patient details-related problems with HIT systems were more likely to affect patient care delivery – more than half of the incidents resulted in patient-related outcomes, namely patient inconvenience and patient harm, including disease risks, severe health deterioration, injury, and even patient death. Incidents associated with patient details can cause deleterious effects; therefore, characterizing them should be a routine part of clinical practice to improve the constantly changing healthcare system.

**Author Summary:** The rapid advances in HIT systems have made healthcare a truly complex socio-technical system than ever before. No matter what changes are introduced in healthcare, new, unforeseen problems always arise. Our research focuses on improving the already existing HIT systems and the care delivery around those systems by solving the clinical problems we encounter in our day-to-day clinical practice rather than building new technologies. The foundation builds on collecting and analyzing incident reports to illuminate the current challenges of Swedish digital healthcare systems and provide a basis for preventive and corrective strategies, thus improving clinical practice. Although a host of mainly technical problems was expected, around two-thirds of the issues were identified to be generated by failures due to human factors. Therefore, several strategies to mitigate these risks can be implemented, such as training healthcare professionals before integrating new HIT systems and designing out the “error-prone features”. Our study provided insight into patient information/documentation-related problems associated with HIT systems and how human and technical factors affect patient care delivery. The analyses may also help the reporters and analysts regarding where preventive and corrective strategies should be addressed to improve the constantly changing healthcare system.

## Introduction

It is evident that Health Information Technology (HIT)-related interventions create viable and timely opportunities to improve accuracy and efficiency in modern medicine (1–3). However, a survey of a nationally representative sample of medical group practices in the US suggested that adopting HIT, such as Electronic Health Record (EHR), is slow and complex and requires a great deal of support (4).

There is ample evidence that patient details can go awry; for example, inflexible electronic forms can result in incorrect orders (5), inaccurate medication requests from a medication ordering system (6), or lost patient data in the EHR system (7). Data entry errors caused by the user can result in incorrect or outdated information remaining in the system and reproducing the same error at several stages of the procedure (8, 9). Healthcare professionals can also delay patient information entry due to their busy schedules or frequent interruptions, resulting in outdated or incomplete information for a more extended period (9). ‘Patient details’, in this context, refer to information-and-documentation-related features of the patient, i.e., any patient information in a healthcare facility is recorded as a document, either in paper or electronic form. For instance, patient demographics and clinical outcomes are usually stored electronically in the health or medical record. Therefore, a record of any patient information has been considered patient details throughout this study.

Several barriers were identified in a recent study by Bjerkan et al. that negatively impacted the nursing practices’ documentation process, including individual, social, organizational, and technological factors (10). It was also reported that healthcare professionals found the process of electronic-record documentation to be onerous, as such records contain too much information (11).

Jabin et al. demonstrated in 2019 that HIT-related incidents occur at each step of the medical imaging workflow process and that human and technical factors play a role in problems related to patient details (12). The human factor involves interactions among humans and other elements of a system, optimizing human well-being and overall system achievement (13). On the other hand, the technical factor refers to the attributes of practices and devices/systems that can influence the performance of an organization (14).

The Swedish Medical Products Agency (MPA) aims to deliver, accord, and contribute to improved healthcare in collaboration with the Swedish eHealth Agency and the Swedish Authority for Privacy Protection. The healthcare providers were recommended to strengthen process measurement and provide leadership to reduce the risks associated with HIT systems (15). Moreover, all Swedish county councils have established computerized reporting systems to which any healthcare practitioner can submit incident reports (16).

According to Magrabi et al., incident reports could be one source among a range of information repositories (16). Reports of HIT-related incidents indicate the gap between the expected and empirically-supported HIT advances; therefore, continuous incident reports and analysis could bridge the gap (12, 17–19). An integrated framework for safety, quality, and risk management, including incident management and information system, was proposed by Runciman et al. (20). The concepts and terms were established in the form of the International Classification for Patient Safety (ICPS)—classification of incident reports and measurement of safety (21). The ICPS helps collect and analyze incident reports to understand what went wrong and how it went wrong.

The incident reports generally consist of information regarding the circumstances surrounding the incidents (type of incident), such as what contributed to these events occurring and their outcomes. “Incident type is a descriptive term for a category made up of incidents of a common nature grouped because of shared, agreed features” (21). One incident may be classified into more than one type of issue, for example, information-related issues and documentation-related issues. Information-related issues may reflect on patient information or characteristics, for example, the reason for seeking care, primary diagnosis, and patient status (12, 22). The document-related issues may include any written, typed, drawn, stamped, or printed text or any document where patient information has been entered. Documents may include nursing medical records, protocols or policies, patient labels, stickers, requests, reports, and medical images (12, 22).

According to the ICPS, “contributing factors are the circumstances, actions or influences which are thought to have played a part in the origin or development of an incident or to increase the risk of an incident” (21). It is notable that an incident may have more than one contributing factor, and one incident may be a contributing factor to another incident (a “recursive” model). “Patient outcome relates to the impact upon a patient which is wholly or partially attributable to an incident” (21). On the other hand, “organizational outcomes” refer to the impacts upon an organization which is wholly or partially attributable to an incident” (21). However, readers interested in the conceptual framework, key concepts, terms, and definitions of the ‘classes’ comprising the ICPS may follow a series of articles published by the World Health Organization with the formation of the World Alliance for Patient Safety. Three scientific papers were published in 2009, with the result of the work developed using a two-stage Delphi survey, participated by 300 experts from a range of fields (21, 23, 24).

The thematic analyses and deductive approaches of the ICPS (25) are suitable for analyzing and interpreting HIT incidents in Swedish healthcare. Since little research has been conducted that has focused on issues related to patient details reported in HIT incidents, there is a need for qualitative analysis, both deductive and inductive. This will help explore the challenges related to patient details (information and documentation) that arise in routine clinical practice in Swedish healthcare.

The overall aim of this study was to explore HIT-related incidents and identify patient details-related problems, as well as their association with human and technical factors, using thematic analysis. The study also examines each HIT incident’s contributing factors and outcome using the ICPS. This paper explores the following research questions:

1. What patient details-related issues occur in the routine clinical practice of Swedish healthcare?
2. How are these problems associated with human and technical factors?
3. What are the other contributing factors and outcomes of these patient details-related issues?

## Methods

### Data collection

Initially, a list of 55 participants was made using purposive sampling, covering 21 regions of Sweden, targeting physicians, nurses, medical engineers, and healthcare quality managers. Of the 55 participants contacted, only five responses were received. Due to this low response, additional 19 participants were approached using snowball sampling, of which 15 responses were collected. The incident reports were collected in two formats depending on the availability and accessibility of the participants. The participants were requested either to participate in interviews (written and telephone) and/or provide a set of retrospectively collected incident reports from their local database.

From the 15 responders, 98 incident reports were collected, three of which were excluded either due to lack of adequate information or inability to categorize it as a HIT incident. The final sample of 95 retrospectively collected HIT incident reports from Swedish healthcare was considered for identifying patient details-related problems and their association with human and technical factors. A detailed description of the participant characteristics from each region, the number of incidents collected, and the time interval of collected incidents are presented in Table 1 (26).

**Table 1.**
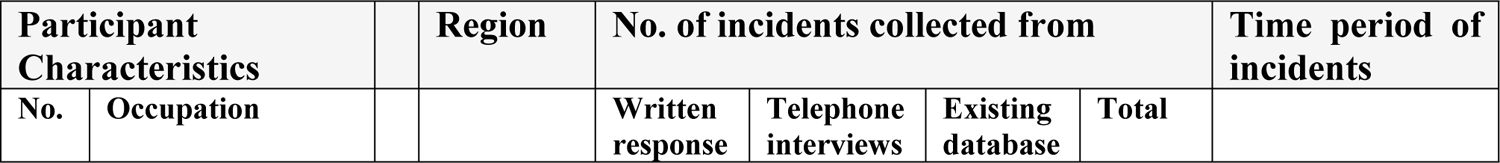
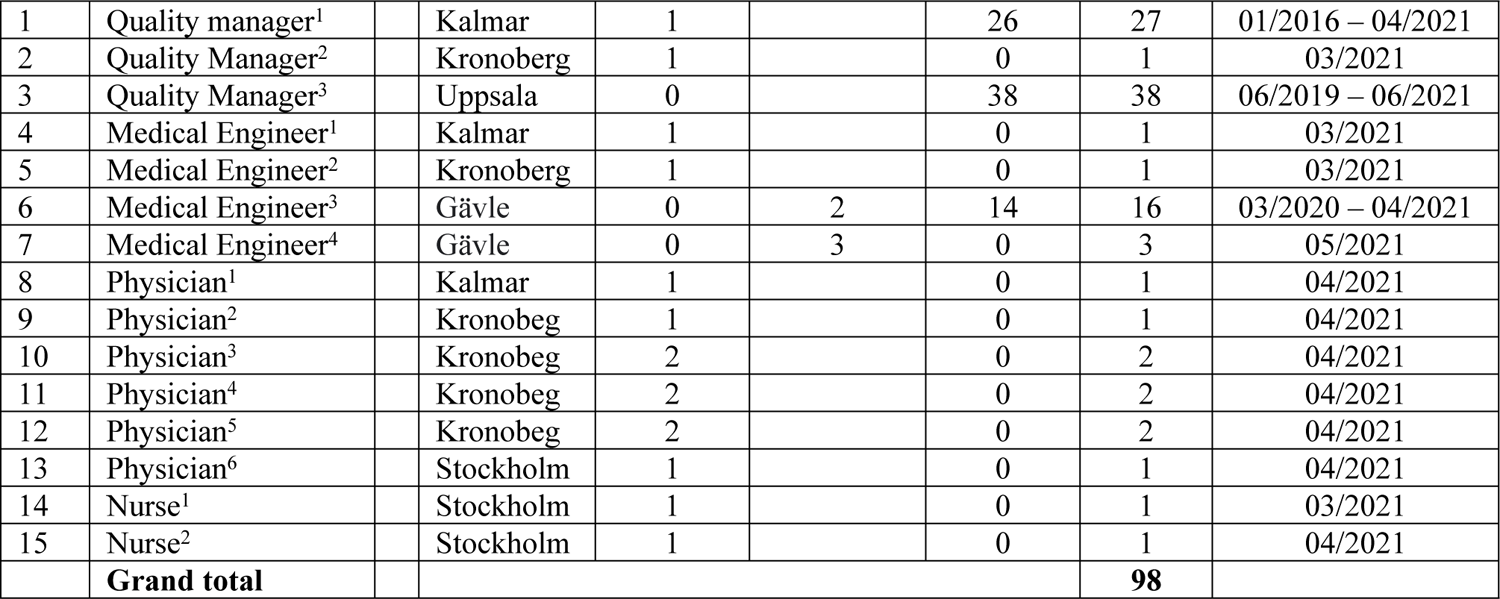
Participant and incident characteristics

### Data analysis

Incident reports (in the form of free-text narratives) were analyzed using both inductive and deductive methods in order to extract detailed information. The inductive approach involved thematic analysis, proposed by Braun and Clarke(27), whereas the deductive included the ICPS. The thematic analysis was used to determine the incidents with patient details from the total sample (n=95). The ICPS was used to identify the contributing factors and outcomes of the incidents. Each identified issue was then characterized by either human or technical factors.

Two coders were involved in data analyses (both deductive and inductive) for verification and reliability of the coding. The primary coder performed the thematic analysis, which the secondary coder verified, and the incident was re-examined in case of any disagreement between the coders. An agreement was reached between the coders through dialogue. The coding of the ICPS (contributing factors and outcomes) was performed by both coders independently. Interrater reliability using kappa score calculation was performed. A consensus was reached in case of any difference of opinion.

## Results

Of 95 included incident reports, 77 were from the existing databases of the local hospitals, and 18 were collected through interviews. Of the 18 incident reports collected via interview, 13 were written responses, and five were telephone interviews. All 95 incidents were aggregated for data analysis. The incidents were reported between January 2016 and May 2021.

Of the total sample (n=95), 90 incidents were associated with problems with patient details using thematic analysis, and 120 issues were identified from these problems (see Table 2). Of 90 incidents, 24 incidents comprised two issues, and three incidents resulted in three problems; however, no particular pattern or common theme was found for the incidents with multiple issues. The 90 incidents fell into three main categories: medical records, e-prescribing, and medical imaging. A fourth category, “other,” included clinical chemistry and psychological treatment, which did not fall into the main three categories. When the 90 HIT incidents were allocated to thematic analysis, 120 issues with patient details were identified, more associated with documentation (n=62, 52%) than information (n=58, 48%) (see Table 2).

**Table 2.** Types of patient detail issues and their association with human and technical factors

| Types of patient detail issues | HF | TF | n | % |
| --- | --- | --- | --- | --- |
| **Medical record-related** |  |  |  |  |
| **Information** |  |  |  |  |
| Incorrect information | 18 | 0 | 18 | 15.00 |
| No/missing/lost information | 12 | 2 | 14 | 11.67 |
| Invalid/ irrelevant/ unknown information | 6 | 1 | 7 | 5.83 |
| Incomplete information | 2 | 0 | 2 | 1.67 |
| **Subtotal (information)** | **38** | **3** | **41** | **34.17** |
| **Documentation** |  |  |  |  |
| No/missing/lost documentation | 0 | 14 | 14 | 11.67 |
| Incorrect documentation | 9 | 1 | 10 | 8.33 |
| Invalid documentation | 6 | 2 | 8 | 6.67 |
| Incomplete documentation | 3 | 0 | 3 | 2.50 |
| **Subtotal (documentation)** | **18** | **17** | **35** | **29.17** |
| **Total (information + documentation)** | **56** | **20** | **76** | **63.33** |

| **e-Prescribing-related** |  |  |  |  |
| --- | --- | --- | --- | --- |
| **Information** |  |  |  |  |
| Incorrect information | 9 | 0 | 9 | 7.50 |
| No/missing/lost information | 3 | 0 | 3 | 2.50 |
| **Subtotal (information)** | **10** | **0** | **12** | **10.00** |
| **Documentation** |  |  |  |  |
| No/missing/lost documentation | 1 | 4 | 5 | 4.17 |
| Invalid documentation | 0 | 4 | 4 | 3.33 |
| Incorrect documentation | 2 | 0 | 2 | 1.67 |
| Incomplete documentation | 0 | 1 | 1 | 0.83 |
| **Subtotal (documentation)** | **3** | **9** | **12** | **10.00** |
| **Total (information + documentation)** | **13** | **9** | **24** | **20.00** |
| **Medical imaging-related** |  |  |  |  |
| **Information** |  |  |  |  |
| No/missing/lost information | 1 | 0 | 1 | 0.83 |
| Incorrect information | 0 | 1 | 1 | 0.83 |
| Invalid information | 1 | 0 | 1 | 0.83 |
| **Subtotal (information)** | **2** | **1** | **3** | **2.50** |
| **Documentation** |  |  |  |  |
| Incorrect documentation | 4 | 1 | 5 | 4.17 |
| No/missing/lost documentation | 0 | 4 | 4 | 3.33 |
| **Subtotal (documentation)** | **4** | **5** | **9** | **7.50** |
| **Total (information + documentation)** | **6** | **6** | **12** | **10.00** |
| **Other** |  |  |  |  |
| **Information** |  |  |  |  |
| No/missing/lost information | 0 | 2 | 2 | 1.67 |
| **Subtotal (information)** | **0** | **2** | **2** | **1.67** |
| **Documentation** |  |  |  |  |
| No/missing/lost documentation | 0 | 2 | 2 | 1.67 |
| Delayed documentation | 0 | 2 | 2 | 1.67 |
| Incorrect documentation | 0 | 1 | 1 | 0.83 |
| Invalid documentation | 0 | 1 | 1 | 0.83 |
| **Subtotal (documentation)** | **0** | **6** | **6** | **5.00** |
| **Total (information + documentation)** | **0** | **8** | **8** | **6.67** |
| **Grand Total (information + documentation)** | **77** | **43** | **120** |  |

HF = Human factor; TF = Technical factor

The problems were also classified to check for association with either human or technical factors. Of 120 issues identified, 77 were determined to be affected by human factors (64%), and the remaining 43 by technical factors (36%). Most problems were contributed to by human factors (n=77; 64%), and the rest by technical factors (n=44; 36%). Within these four groups, each issue was further categorized as either an information-related or documentation-related issue (see Table 2).

Interrater reliability for the outcomes was к (weighted) = 0.89 (p < 0·001, 95% CI 0·81–0·98), and for the contributing factors was к (weighted) = 0.87 (p<0.001, 95% CI 0.78–0.96).

NB: It was assumed that all incidents would be associated with patient details; however, five incident descriptions did not contain patient-related information or documentation. For example, the narration of an incident described a service for automatic drug dispensing for which the service unit failed to cut the seam between medicine bags.

### Information-related issues

Of the 120 issues, 48% (n=58) comprised information-related problems (see Table 3). An example of an information-related issue may include healthcare professionals omitting patient information or writing inaccuracies in the medical record, such as blood pressure medication.

**Table 3.** Types of information-related issues and their association with human and technical factors

| Medical record-related information |  |  |  |
| --- | --- | --- | --- |
| **Incorrect information** |  |  |  |
| Incorrect patient status of care | HF | 5 | 8.62 |
| Incorrect information **on** diagnosis/treatment/care | HF | 5 | 8.62 |
| Incorrect personal/demographic data | HF | 4 | 6.90 |
| Incorrect information about appointment/booking | HF | 3 | 5.17 |
| Incorrect examination/test data | HF | 1 | 1.72 |
| **No/missing/lost information** |  |  |  |
| No/Missing personal/demographic data | HF | 5 | 8.62 |
| No/missing information about appointment/booking | HF | 3 | 5.17 |
| Missing information about student's health status | HF | 2 | 3.45 |
| No/Missing examination/test data | HF | 2 | 3.45 |
| *No retrieval of personal/ demographic data* | *TF* | 2 | 3.45 |
| **Invalid/ irrelevant/ outdated information** |  |  |  |
| Invalid information of diagnosis/treatment/care | HF | 4 | 6.90 |
| Irrelevant personal information | HF | 1 | 1.72 |
| *Outdated personal data* | *TF* | 1 | 1.72 |
| Unknown examination data | HF | 1 | 1.72 |
| **Incomplete information** |  |  |  |
| Incomplete clinical history | HF | 2 | 3.45 |
| **Total** |  | **41** | **70.69** |
| e-Prescribing-related information |  |  |  |
| **Incorrect information** |  |  |  |
| Incorrect dispensing time of medication | HF | 4 | 6.90 |
| Incorrect dose of medication | HF | 4 | 6.90 |
| Incorrect drugs prescribed | HF | 1 | 1.72 |
| **No/missing/lost information** |  |  |  |
| Missing information about hypersensitivity (drugs/foods) | HF | 3 | 5.17 |
| **Total** |  | **12** | **20.69** |
| Medical imaging-related information |  |  |  |
| **No/missing/lost information** |  |  |  |
| Missing concluding information (summary) | HF | 1 | 1.72 |
| **Incorrect information** |  |  |  |
| *Incorrect display of information in medical image* | *TF* | 1 | 1.72 |
| **Invalid/ irrelevant/ outdated information** |  |  |  |
| Invalid (multiple) tumor iso-centre | HF | 1 | 1.72 |
| **Total** |  | **3** | **5.17** |
| Other information |  |  |  |
| **No/missing/lost information** |  |  |  |
| *No/missing administrative information* | *TF* | 2 | 3.45 |
| **Total** |  | **2** | **3.45** |
| **Grand total** |  | **58** |  |

HF = Human factor (non-italics); TF = Technical factor (Italics)

Of the 58 patient information-related problems, more than two-thirds involved medical records (n=41; 71%) (see Table 3). Of these 41 issues, 38 were associated with human factors and three with technical factors (see Table 2). For instance, the physician prescribing an incorrect medication was considered to be associated with the human factor, whereas doctors and pharmacists did not have the same view of the medication list attributed to technical factor-related issues. The most common problem with medical records was “incorrect information” (n=18), and all such problems were attributed to human factors.

A single patient information-related problem may include more than one type of information. For example, incorrect medical record information may encompass several types of demographic or personal information, including name, social security number, or sex. A list of the different types of information is presented in Table 4.

**Table 4.** Type of information involved in the information-related issues

| Type of information | n | % |
| --- | --- | --- |
| **Clinical information** |  |  |
| Diagnostic information | 6 | 6.74 |
| Patient status | 4 | 4.49 |
| Care measures | 3 | 3.37 |
| Care plan | 2 | 2.25 |
| Vaccination registration | 2 | 2.25 |
| Alcohol history | 1 | 1.12 |
| Investigation | 1 | 1.12 |
| Intervention (type) | 1 | 1.12 |
| **Total** | **20** | **22.47** |
| **Demographic/personal information** |  |  |
| Patient name | 7 | 7.87 |
| Patient identity | 3 | 3.37 |
| Social security number | 3 | 3.37 |
| Sex | 3 | 3.37 |
| Student guardians information | 2 | 2.25 |
| Student class affiliation | 1 | 1.12 |
| **Total** | **19** | **21.35** |
| **Medication-related information** |  |  |
| Drug dose | 5 | 5.62 |
| Medicine | 4 | 4.49 |
| Drug dispensing information | 3 | 3.37 |
| **Total** | **12** | **13.48** |
| **Requests/referral related** |  |  |
| Appointments | 4 | 4.49 |
| Electronic order | 3 | 3.37 |
| *Referral* | 3 | 3.37 |
| Examination request | 1 | 1.12 |
| **Total** | **12** | **13.48** |
| **Pathophysiological (disease)-related information** |  |  |
| *Hypersensitivity* | 4 | 4.49 |
| Aortic rupture | 1 | 1.12 |
| Emergency medical care | 1 | 1.12 |
| Cardiac arrest event | 1 | 1.12 |
| Bleeding | 1 | 1.12 |
| Suicide | 1 | 1.12 |
| **Total** | **9** | 10.11 |
| **Personal measures** |  |  |
| Weight | 2 | 2.25 |
| Back | 1 | 1.12 |
| Hearing | 1 | 1.12 |
| Height | 1 | 1.12 |
| Vision | 1 | 1.12 |
| **Total** | **6** | 6.74 |
| **Examination/Test related** |  |  |
| Examination items | 1 | 1.12 |
| Sampling time | 1 | 1.12 |
| Sampling responses | 1 | 1.12 |
| Test date | 1 | 1.12 |
| Type of examination | 1 | 1.12 |
| **Total** | **5** | 5.62 |
| **Administration related** |  |  |
| Patient bed | 1 | 1.12 |
| Staff categories | 1 | 1.12 |
| Term of benefit | 1 | 1.12 |
| **Total** | **3** | 3.37 |
| **Medical imaging** |  |  |
| Tumor isocentre central | 1 | 1.12 |
| Tooth graphic | 1 | 1.12 |
| **Total** | **2** | 2.25 |
| **Communication-related** |  |  |
| Health follow-up | 1 | 1.12 |
| **Total** | **1** | 1.12 |
| **Grand Total** | **89** |  |

Patient details-related issues associated with patient information fell into four categories: incorrect information, no/missing/lost information, invalid/irrelevant/outdated information, or incomplete information. There were consistent indications of the things going wrong in which human failures played a major role. Among information-related issues, human factors manifested “incorrect information” (27 of 28) and “no/missing/lost information” (16 of 20) (see Table 5).

**Table 5.** Finite categories of information-related issues and their association with human and technical factors

| Finite categories of information-related issues | HF | TF | n | % |
| --- | --- | --- | --- | --- |
| Incorrect information | 27 | 1 | 28 | 48.28 |
| No/missing/lost information | 16 | 4 | 20 | 34.48 |
| Invalid/ irrelevant/ outdated information | 7 | 1 | 8 | 13.79 |
| Incomplete information | 2 | 0 | 2 | 3.45 |
| **Total** | **52** | **6** | **58** |  |

HF = Human factor; TF = Technical factor

### Documentation-related issues

Of the 120 problems, 52% (n=62) were document-related issues (see Table 6). An illustration of documentation-related issues may involve multiple imaging requests that were required to be written in paper form because of a malfunction of the radiology ordering system.

**Table 6.** Types of documentation-related issues and their association with human and technical factors

| Types of documentation-related issues | HF/TF | n | % |
| --- | --- | --- | --- |
| **Medical records-related documentation** |  |  |  |
| **No/missing/lost documentation** |  |  |  |
| *Missing patient record* | *TF* | 5 | 8.06 |
| *No/Missing sick/medical certificate* | *TF* | 3 | 4.84 |
| *No/missing request/order/appointment form* | *TF* | 3 | 4.84 |
| *Missing patient report* | *TF* | 2 | 3.23 |
| *Missing waiting list entry* | *TF* | 1 | 1.61 |
| **Incorrect documentation** |  |  |  |
| Incorrect patient record | HF | 6 | 9.68 |
| Incorrect request/order/appointment form | HF | 3 | 4.84 |
| *Another patient's request form* | *TF* | 1 | 1.61 |
| **Invalid/ outdated documentation** |  |  |  |
| Unauthorized access/process to medical record | HF | 5 | 8.06 |
| *Outdated documentation* | *TF* | 2 | 3.23 |
| Confused medical record | HF | 1 | 1.61 |
| **Incomplete documentation** |  |  |  |
| Incomplete document for care plan | HF | 3 | 4.84 |
| **Total** |  | **35** | **56.45** |
| **e-Prescribing-related documentation** |  |  |  |
| **No/missing/lost documentation** |  |  |  |
| *Missing prescription* | *TF* | 3 | 4.84 |
| *No display of "terms of benefit"* | *TF* | 1 | 1.61 |
| No medical prescription sent | HF | 1 | 1.61 |
| **Invalid/ outdated documentation** |  |  |  |
| *Outdated prescription* | *TF* | 2 | 3.23 |
| *Different drug list* | *TF* | 2 | 3.23 |
| **Incorrect documentation** |  |  |  |
| Another patient's prescription | HF | 2 | 3.23 |
| **Incomplete documentation** |  |  |  |
| *Incomplete display of prescription* | *TF* | 1 | 1.61 |
| **Total** |  | **12** | **19.35** |
| **Medical Imaging-related documentation** |  |  |  |
| **Incorrect documentation** |  |  |  |
| Incorrect medical image obtained/transferred | HF | 4 | 6.45 |
| *Incorrect generation of X-ray referral* | *TF* | 1 | 1.61 |
| **No/missing/lost documentation** |  |  |  |
| *No retrieval of medical image* | *TF* | 3 | 4.84 |
| *No transfer of image* | *TF* | 1 | 1.61 |
| **Total** |  | **9** | **14.52** |
| **Other documentation** |  |  |  |
| **No/missing/lost documentation** |  |  |  |
| *No message/chat* | *TF* | 2 | 3.23 |
| **Delayed documentation** |  |  |  |
| *Delayed delivery of the message* | *TF* | 2 | 3.23 |
| **Incorrect documentation** |  |  |  |
| *Incorrect message delivered* | *TF* | 1 | 1.61 |
| **Invalid documentation** |  |  |  |
| *Different alarm logs* | *TF* | 1 | 1.61 |
| **Total** |  | **6** | **9.68** |
| **Grand Total** |  | **62** |  |

HF = Human factor (non-italics); TF = Technical factor (Italics)

Of the 62 documentation-related problems, more than half were associated with medical records (n=35; 56%) (see Table 4). Of these 35 issues, 18 were attributed to human factors and 17 to technical factors (see Table 6). The most common issue with medical records was “no/lost/missing documentation” (n=14), of which all were associated with technical factors. A list of document types is presented in Table 7, of which fewer than half comprised clinical documents (n=24; 44%).

**Table 7.** Type of documentation involved in the documentation-related issues

| **Type of document involved** | **n** | **%** |
| --- | --- | --- |
| **Clinical document** |  |  |
| Medical record | 11 | 20.00 |
| Patient report | 5 | 9.09 |
| Certificates (sick) | 4 | 7.27 |
| Patient meeting documentation | 1 | 1.82 |
| Waiting list | 1 | 1.82 |
| Checklist | 1 | 1.82 |
| Treatment template | 1 | 1.82 |
| **Total** | **24** | **43.64** |

| Medication-related document |  |  |
| --- | --- | --- |
| Prescription | 8 | 14.55 |
| Drug lists | 2 | 3.64 |
| **Total** | **10** | 18.18 |
| Medical imaging-related document |  |  |
| Medical image | 8 | 14.55 |
| Imaging report | 2 | 3.64 |
| **Total** | **10** | 18.18 |
| Request/booking form |  |  |
| Request form | 3 | 5.45 |
| Booking form | 2 | 3.64 |
| **Total** | **5** | 9.09 |
| Communication-related document |  |  |
| Message (chat) | 5 | 9.09 |
| **Total** | **5** | 9.09 |
| Other documents |  |  |
| Alarm logs | 1 | 1.82 |
| **Total** | **1** | 1.82 |
| **Grand Total** | **55** |  |

Documentation-related patient-detail problems fell into five categories: no/missing/lost documentation, incorrect documentation, invalid/outdated documentation, incomplete information, or delayed documentation. Among these, human factors predominated in the causation of “incorrect documentation” (15 of 18), while technical factors predominated in “no/missing/lost documentation” (24 of 25) (see Table 8).

**Table 8.** Finite categories of documentation-related issues and their association with human and technical factors

| Finite categories of documentation-related issues | HF | TF | n | % |
| --- | --- | --- | --- | --- |
| No/missing/lost documentation | 1 | 24 | 25 | 40.32 |
| Incorrect documentation | 15 | 3 | 18 | 29.03 |
| Invalid/ outdated documentation | 6 | 7 | 13 | 20.97 |
| Incomplete documentation | 3 | 1 | 4 | 6.45 |
| Delayed documentation | 0 | 2 | 2 | 3.23 |
| **Total** | **25** | **37** | **62** | **51.67** |

HF = Human factor; TF = Technical factor

### Contributing factors

Te ICPS was used to capture detailed information about the types of contributing factors associated with those 90 incidents involving patient details-related issues. Of these 90 incidents, 87 contributing factors were identified, of which fewer than half (45%) comprised “medical device/system” factors (n=39) (see Table 9).

**Table 9.** Types of contributing factors involved in the patient details-related issues

| **Contributing factors** | **n** | **%** |
| --- | --- | --- |
| **Medical device/ system factor** |  |  |
| Device or system usability | 16 | 18.39 |
| Device or system not working/slow/ failed | 11 | 12.64 |
| Device or system suitability for purpose | 9 | 10.34 |
| Device or system unavailable/inaccessible | 2 | 2.30 |
| Device or system unfamiliar | 1 | 1.15 |
| **Subtotal** | **39** | **44.83** |
| **Documentation factor** |  |  |
| Missing/unavailable documentation | 7 | 8.05 |
| Unclear/ambiguous/ duplicated documentation | 4 | 4.60 |
| Breach of privacy | 4 | 4.60 |
| Inadequate/ incomplete documentation | 2 | 2.30 |
| **Subtotal** | **17** | **19.54** |
| **Staff factor** |  |  |
| Unknown/ Not clear | 10 | 11.49 |
| Inattention/distraction/negligence | 3 | 3.45 |
| Knowledge/skills/awareness | 3 | 3.45 |
| Fail to carry out duty | 1 | 1.15 |
| **Subtotal** | **17** | **19.54** |
| **Communication factor** |  |  |
| Not conducted | 6 | 6.90 |
| Inaccurate information communicated | 3 | 3.45 |
| Incomplete | 1 | 1.15 |
| **Subtotal** | **10** | **11.49** |
| **Policy/guideline-related factor** |  |  |
| Policy/guideline not followed | 2 | 2.30 |
| **Subtotal** | **2** | **2.30** |
| **Patient factor** |  |  |
| Inattention/distraction/negligence | 1 | 1.15 |
| Knowledge/skills/awareness | 1 | 1.15 |
| **Subtotal** | **2** | **2.30** |
| **Total** | **87** |  |

Among medical device/system-related factors, “device/system usability” (n=16) was most common; for instance, the computer system malfunctioned at the beginning of imaging because the imaging modality was turned on in haste. Both documentation (n=17) and staff (n=17) each contributed to one-fifth (20%) of the total factors (see Table 9). Documentation factors were mainly associated with “missing/unavailable documentation” (n=7); for example, a patient summary could not be retrieved because the patient record was missing.

However, staff-related factors were largely unclear or not known due to insufficient narration of the incident descriptions (see Table 9). In these cases, it was quite clear that healthcare staff contributed to an incident, but information about the type of contributing factor was lacking. For example, a report indicated that staff omitted information or wrote inaccuracies in the medical record, but no indication was given of their reason for doing so.

Regarding “medical device/system,” the most common factor was “device or system usability” (n=16; 18%), indicating that users or healthcare staff had difficulty using the system, which contributed to the incident. For example, a user could not enter complete patient prescription details because benefit terms were not displayed in the intended context. 11% of incidents comprised the communication factor, of which communication was “not conducted” in six cases (see Table 9).

Moreover, “medical device/system” and “documentation” were the most common contributing factors, especially when the incidents themselves were HIT system issues (in nature) and involved documentation-related problems (through thematic analysis). This phenomenon confirms the recursive nature of errors and the frequent correlation between types of problems and specific contributing factors. Therefore, the phenomenon may be considered an issue, a contributing factor, or both based on the incident’s features or characteristics in that specific context.

### Outcomes

The outcomes of all 90 incidents were identified using the ICPS, and each incident was assigned one outcome. The outcomes were broadly classified into two categories, namely, patient-related, comprising patient inconvenience and patient harm, or staff/organization-related. Of 90 incidents, more than half (59%) comprised patient-related outcomes (n=53)—patient inconvenience (n=42; 47%) and patient harm (n=11; 12%). The remaining 41% (n=37) comprised staff or organization-related outcomes (see Table 10).

**Table 10.** Types of outcomes resulting from the patient details-related issues

| Outcomes | N | % |
| --- | --- | --- |
| **Patient-related outcomes** |  |  |
| **Patient inconvenience** |  |  |
| Delays in management diagnosis/procedure/treatment | 17 | 18.89 |
| Unnecessary treatment | 9 | 10.00 |
| Patient dissatisfaction | 8 | 8.89 |
| Repeated or additional diagnosis/procedure/treatment | 5 | 5.56 |
| Financial implications | 3 | 3.33 |
| **Subtotal** | **42** | **46.67** |
| **Patient harm** |  |  |
| Pathophysiological disease related | 6 | 6.67 |
| Injury | 4 | 4.44 |
| Death | 1 | 1.11 |
| **Subtotal** | **11** | **12.22** |
| **Total** | **53** | **58.89** |
| **Staff or organization-related outcomes** |  |  |
| More system/service/resource used | 14 | 15.56 |
| Delays in using facilities/service/system | 10 | 11.11 |
| Increased documentation | 7 | 7.78 |
| Phone calls/review/follow-up | 6 | 6.67 |
| **Subtotal** | **37** | **41.11** |
| **Total** | **37** | **41.11** |
| **Grand total** | **90** |  |

Within patient inconvenience, delays in care management (diagnosis/procedure/treatment) accounted for 19% (n=17) of outcomes. Such delays included investigation of the patient treatment plan, care measures, acquisition of images, or medication management. Unnecessary treatment (n=9; 10%) included patient treatment that was not required; for example, patient treatment with blood pressure medication despite any such symptoms or imaging with unnecessary radiation dose. Patient dissatisfaction (n=8; 9%) was expressed either in informal complaints or expressions of dissatisfaction, including decreased confidence in care delivery, suspicions of unauthorized medical records access, or concerns about the competence of the care providers. Repeated or additional diagnosis/procedure/treatment (n=5; 6%) was performed when the healthcare professionals found that the wrong patient was treated/imaged or that the right patient underwent the wrong treatment or examination. These problems caused the same patient to undergo the same imaging, examination, or treatment twice. Three cases of financial implications for patients were identified, including denial of payment or failure to pay compensation or sickness benefit (see Table 10).

Six cases resulted in pathophysiological disease-related harm to the patient, including risks of disease or severe deterioration of health due to wrong medication. These comprised four cases of serious injury and one patient death. However, no further information regarding the injury was reported, and the death was not reported to be directly caused by the incident (see Table 10).

Of the staff or organization-related outcomes, 16% (n=14) resulted in more equipment/services/resources being used in the form of similar systems or USB sticks, manual extraction/entry of data, or other healthcare professionals, including medical engineers or IT experts. 11% of outcomes resulted in delays in using facilities/service/systems, for example, storage server access delays due to system shutdown; 8% led to increased documentation, such as rewriting imaging orders in paper form; and the remaining 7% resulted in phone calls/review/follow-up, such as contacting other staff involved in the same treatment (see Table 10).

## Discussion

In the last three decades, healthcare quality and safety have been on the agenda because the healthcare system can harm patients; for example, a single large-scale event may even affect multiple patients’ care management (26). Even though HIT has improved efficiency, its design, use, and implementation can negatively impact patient care and safety (12, 28). The Joint Commission that accredits healthcare organizations in the US suggested that diligent adherence to protocol, cross-checking HIT systems’ information and documents, and the use of interpreters for foreign patients could potentially detect and mitigate such problems (29).

A report in Australia suggests that patient information and documentation issues were significant modifiable risks in medical imaging, nuclear medicine, radiation therapy, and oral surgery. These risks persisted even after developing and implementing the Correct Patient, Correct Site, and Correct Procedure (3Cs) Protocol in 2004 (30). However, caution is advised since the daily average volume of medical exams read by radiologists has increased sevenfold in the last seven years. As thousands of patients are processed, transported, treated, and examined by hundreds of healthcare staff in the day-to-day clinical routine, the risks for such failures are enormous (30). The report further indicated that the factors contributing to such incidents include “heavy workload” and “policies/guidelines not followed”, suggesting that healthcare professionals do not receive enough time to follow the protocol (30).

### Human versus technical factors

Healthcare is a complex sociotechnical system in which various human factors, including behavior, performance, and culture, play a vital role in building an intimate relationship with the HIT systems. These factors can improve healthcare quality and safety or cause harm and disrupt healthcare processes (31). Even though many technical issues and failures were identified in the reports, we did not expect that more than half of the issues would be caused by human factors (n=77; 64%). However, the incident reports did not contain adequate information to explore further any connection with human behavior, performance, and culture.

Despite the analysis providing no indication of the absolute frequencies of these issues, it does assures the fact that human errors play a vital role in HIT incidents (32). The advantage of systems is that they can be incrementally improved, while the errors of human users are inevitable and remain an inherent part of the complex sociotechnical healthcare system. “Whilst system issues can be progressively “designed out”, it would seem that in the meantime, the rapid availability of experts to diagnose and apply a digital solution to such problems would be highly desirable” (17). Therefore, more considerable thought should be placed on the HIT systems to be designed to prevent specific issues from occurring. This can also be backed up by observational and ethnographic studies that would prevent the occurrence of issues such as those listed throughout this study (28, 32).

### Humans as the weak link, and the need for training for healthcare professionals

A review of 436 HIT incident reports by Jabin et al. reported that human factors are inevitable in the genesis of over 58% of the issues in most complex sociotechnical systems (12). Magrabi et al., in a review of 850 incidents, summarized that human factors were responsible for patient harm four times more often than technical factors (18).

Human error is difficult (if not impossible) to prevent because neither seniority nor experience offers immunity (33). An error occurs through various unintended and unknown cognitive mechanisms beyond human control (34). Even though using the “forcing function” was suggested to prevent such human errors by Norman (34), it would cause “over-proceduralization” that may detract from surveillance and situational awareness. Therefore, it would rather be suitable to design the system in such a way as to prevent incidents from occurring, as well as provide training to system users (17).

Healthcare professionals are seldom provided with sufficient training or education for the proper use and operation of HIT systems, manifesting as a lack of proficiency in handling them in healthcare settings (35). For example, a review of 436 HIT incidents by Jabin et al. suggests that system integration and software update-related issues do contribute to human error, causing incorrect entry of patient information and workflow disruptions (17).

Since human error remains an enduring part of the complex healthcare system, it is critically essential to establish an ongoing training process for healthcare professionals in connection with vendors (17). The Joint Commission suggested that healthcare professionals, as the major contributor to HIT-related problems, should be adequately trained (29). Therefore, training healthcare professionals before integrating HIT systems and software updates, and preparing them for unexpected system failures, will result in better use of the HIT systems and greater user satisfaction, thus mitigating the risks of human problems.

However, professional development to acquire HIT-related skills and competencies can affect patient safety as it hinders clinicians from regular routine practice, such as patient care activities (36). One of the effective approaches to overcome this barrier is to combine conventional classroom training with simulation-based training (37). This approach is useful for novice user but not for experienced participants who may intend to refrain from any additional training (37). Another approach is to set aside adequate paid time for balancing the training and their day-to-day clinical practice (17). These approaches can also be supplemented by basic health literacy programs, which may involve patients in managing their own care (38). For example, a defense mechanism in the general healthcare system can be improved by identifying incorrect prescriptions by means of providing basic health literacy to patients (39).

Therefore, training healthcare professionals before integrating HIT systems and software updates, and preparing them for unexpected system failures, will result in better use of the HIT systems and greater user satisfaction, thus mitigating the risks of human problems.

### Lack of human-centered design of HIT systems

Even if it is called the “human factor” or “human errors”, it should be clarified that often it is not individuals who are to be blamed. Rather, it is the complex systems the healthcare professionals work in that are not sufficiently designed (40). Although there was a separation of the human factors from the technical in this study, humans constantly interact with multiple other systems or elements when performing their jobs in complex systems. These include people, job tasks, technology, physical and social environments, the organization of work, and external issues such as regulation and research findings (41, 42). Often, systemic influences and reasons that are either unknown/or unknowable fall under the category of system design–human interaction issues (43, 44).

The concept of ‘human error’ as it applies in complex sociotechnical systems, combined with the lack of human-centered HIT design, causes the system to be unsafe and suffer from usability issues (45). The great majority of data reported, therefore, are, in fact, human factors-related issues. Therefore, blaming the end-user of these systems as contributors to these incidents without extensive evidence occurring in day-to-day clinical practice is not appropriate.

Human error can potentially be mitigated by designing systems to prevent incidents from occurring and designing out the “error-prone features”. With considerable thought and ingenuity, the National Health Service (NHS) and the US National Institute of Standards and Technology (NIST) developed and published guidelines and standards for interface design for clinical user interface (45) and EMR usability (46). The HIT industries and national standardization bodies should step in for the design and development of well-established guidelines and standards for safety-critical software. These guidelines and measures should be established based on coveted and suitable working procedures, maintained by national standardization bodies, and backed by government authorities (47).

### Disruptions in the clinical workflow and the need for a holistic view of healthcare

In this study, approximately 41% of cases had a staff/organization-related outcome. There was a clear indication that workflow disruptions resulted in additional system/service/resource use and delays in using facilities/service/systems. More than one-third of problems in this study were associated with incorrect patient information or documentation (n=46; 38%). These events caused several risks to patients, such as increased radiation risks, unnecessary procedures, or delays in obtaining correct procedures or medication. Such delays in procedure further delayed diagnosis, treatment initiation, treatment impact monitoring, and decisions regarding future treatment options (continuation, discontinuation, or change in treatment). Once an incorrect piece of information or document is introduced into the system, an “automation bias” tends to consider it correct (48).

Over the decades, health informatics researchers have been studying the effect of patient information and documentation-related problems that affect the clinical workflow. A review of 149 HIT incidents by Warm and Edwards in 2012 reported that around 34% of the total sample was patient information-related issues, which were categorized into information output (n=25; 16%), information transfer (n=7; 5%), and information input (n=19; 13%) (49). Other studies reported that the quality of the care delivery was compromised by less-than-optimal care, or the risks to patient safety were caused by things going wrong with care delivery (17, 50). Some of the negative impacts include delayed procedures (5), confusion about the patient treatments (51), inappropriate decision-making based on incorrect or outdated information (52), and patient harm (51, 53).

A holistic view of the healthcare workflow is necessary to understand and identify the risks. This could include an assessment of risks among various healthcare departments, such as medical imaging, emergency departments, theatres, and Intensive Care Units (ICU). This risk assessment should be accompanied by the ongoing development of new effective strategies to mitigate risk. For example, Jabin et al. reported that failures related to patient information or documentation can occur at any stage, such as clinical consultation to clinical action with particular focus on the medical imaging workflow process (12). This is true even in our study of the incidents associated with patient identification issues, irrespective of the process workflow in medical imaging or e-prescribing. However, the present study did not consider clinical workflow stages or their association with patient detail issues. Therefore, it is essential to examine different types of patient information and documentation problems, their causation, and their effects on the clinical workflow. This can further help develop the workflow and design solutions addressing particular issues for each healthcare department.

### Strengths and limitations of this study

Incident reports are voluntary, subject to bias, and self-reported. The low response rate was due to the ongoing COVID-19 pandemic, which resulted in a smaller sample than planned. The reporters were not experts in HIT (54), as there needed to be more accurate in the reported narrative texts, making it impossible to categorize three incidents. For example, an incident was assigned at least one contributing factor; however, it was impossible to classify three incidents to identify even one contributing factor due to insufficient incident description. Therefore, 12% of staff factor-related incidents were categorized as “unknown/unclear” (see Table 7). The inadequacy in the reports also affected the categorization of information-/documentation-related issues; for example, “no” information/documentation was merged with “missing/lost” information/documentation. In addition, most of the issues affected by the “staff factor” could not be identified for the same reason—insufficient narrative texts in the report.

It was a limited data source, and thus it is assumed that incidents were not detected in many cases and not reported even if they were detected (55). Moreover, the total number or frequency of events (failures) can never be compared to successful actions that occur in healthcare. Therefore, the analyses cannot manifest the absolute frequency of the issues, contributing factors, or the identified outcomes. However, the numbers and frequency do provide a salient sense of HIT issues that occur in day-to-day clinical practice and demonstrate the harmful role of human error in patient details-related problems.

The incident reports were analyzed using thematic analysis and classified using the ICPS by an expert analyst who previously investigated, analyzed, and classified a large set of incident reports—around 5,000 medical imaging incident reports in Australia. The secondary coder was extensively trained to classify the incidents using the ICPS. Also, the ICPS was initially developed without considering the HIT system; therefore, a slight modification of the contributing factor nomenclature “medical device/system” has been considered for this study. Moreover, a combination of both deductive and inductive approaches made it possible to extract information that may not be evident using any one method of analysis. Moreover, the recursive model of the descriptions of errors and how they occurred constitutes a measure of internal validation for the incident reporting and classification process.

The collection of incident reports ranged over a significant period, validating the feasibility of monitoring incidents regularly. Therefore, new features of the existing problems may have emerged, and new, unforeseen, and unprecedented issues may have been identified. The issues identified in this study are compliant with amelioration and mitigation through a systemic approach and rigorous research, such as providing training to healthcare professionals, vigilant system design and implementation, and redesigning the clinical workflow. The issues can be mitigated by permitting timely application of preventive and corrective strategies at systemic and local levels.

Even though minimal studies on incident analysis have been published, Sweden currently has a comprehensive program on medical device and HIT systems managed by the MPA to ensure quality improvement in Swedish healthcare. Therefore, the results obtained, i.e., the broader categories of the issues, are similar to those previously identified in Australia (32) and the UK (49). This means the lessons learned may be suitable and applicable elsewhere to maintain risk management standards.

## Conclusion

This study provided insight into patient information/documentation-related problems vis-à-vis HIT and how human and technical factors affect patient care delivery. The deductive and inductive approaches analyses provided useful context to the reporters and analysts regarding where preventive and corrective strategies should be addressed. Therefore, characterizing such HIT incidents and identifying patient details-related problems should be a routine part of clinical practice to improve the constantly changing healthcare system.

## Data Availability

We wish we could share the data. Unfortunately, we cannot share this data since we do not have permission to do so. However, we would sincerely request you grant us the exemption of data availability

## Acknowledgment

The authors wish to thank Sofia Backåberg and Pauline Johansson, Senior Lecturers at Linnaeus University for their help and support.

## Notes

### Competing Interest Statement

The authors have declared no competing interest.

### Funding Statement

The author(s) received no specific funding for this work.

### Author Declarations

Ethical advice (Dnr 701-2021) was received from the Ethical Advisory Board in South East Sweden on 4th March 2021, prior to this study. No personal or sensitive information was collected. The datasets of incident reports were filtered to deidentify any personal or sensitive information by the respective management before providing them to us. The feedback from the committee was: The ethics committee SouthEast has taken note of this application for the planned study. The committee sees no ethical difficulties with it being carried out as planned student work with registration and/or publication in DIVA.

